# KMSubtraction: Reconstruction of unreported subgroup survival data utilizing published Kaplan-Meier survival curves

**DOI:** 10.1101/2021.09.04.21263111

**Authors:** Joseph J. Zhao, Nicholas L. Syn, Benjamin Kye Jyn Tan, Dominic Wei Ting Yap, Chong Boon Teo, Yiong Huak Chan, Raghav Sundar

**Affiliations:** Yong Loo Lin School of Medicine, National University of Singapore, Singapore; Biostatistics Unit, Yong Loo Lin School of Medicine, National University of Singapore, Singapore; Department of Haematology-Oncology, National University Health System, Singapore; Cancer and Stem Cell Biology Program, Duke-NUS Medical School, Singapore; The N.1 Institute for Health, National University of Singapore, Singapore; Singapore Gastric Cancer Consortium

## Abstract

**BACKGROUND:** Data from certain subgroups of clinical interest may not be presented in primary manuscripts or conference abstract presentations. In an effort to enable secondary data analyses, we propose a workflow to retrieve unreported subgroup survival data from published Kaplan-Meier (KM) curves.

**METHODS:** We developed *KMSubtraction*, an R-package that retrieves patients from unreported subgroups by matching participants on KM curves of the overall cohort to participants on KM curves of a known subgroup with follow-up time. By excluding matched patients, the opposing unreported subgroup may be retrieved. Reproducibility and limits of error of the *KMSubtraction* workflow were assessed by comparing unmatched patients against the original survival data of subgroups from published datasets and simulations. Monte Carlo simulations were utilized to evaluate the effect of the reported subgroup proportion, missing data, censorship proportion in the overall and subgroup cohort, sample size and number-at-risk table intervals on the limits of error of *KMSubtraction*. 3 matching algorithms were explored – minimal cost bipartite matching, Mahalanobis distance matching, and nearest neighbor matching by logistic regression.

**RESULTS:** The validation exercise found no material systematic error and demonstrates the robustness of *KMSubtraction* in deriving unreported subgroup survival data. Limits of error were small and negligible on marginal Cox proportional hazard models comparing reconstructed and original survival data of unreported subgroups. Extensive Monte Carlo simulations demonstrate that datasets with high reported subgroup proportion (r=0.467, p<0.001), small dataset size (r=-0.374, p<0.001) and high proportion of missing data in the unreported subgroup (r=0.553, p<0.001) were associated with uncertainty are likely to yield high limits of error with KMSubtraction.

**CONCLUSION:** While *KMSubtraction* demonstrates robustness in deriving survival data from unreported subgroups, the implementation of *KMSubtraction* should take into consideration the aforementioned limitations. The limits of error of *KMSubtraction*, as reflected by the mean |ln(HR)| from converged Monte Carlo simulations may guide the interpretation of reconstructed survival data of unreported subgroups.

## BACKGROUND

Advances in secondary data analysis of survival data has been made by Guyot et al.^1^ This enables retrieval of individual patient data (IPD) from reported KM curves.^2,3^ However, this is only amenable for KM curves presented in the original publication. Often, subgroups of interest in clinically negative randomized controlled trials (RCTs) may not be presented in Kaplan-Meier (KM) curves. Where subgroups of interest are not reported (or more commonly presented as a summary statistic in forest plots), IPD retrieval utilizing such algorithms becomes impossible.

In this era of precision medicine, where molecular markers or gene expression levels are increasingly available to clinicians, treatment should ideally be rendered to the patient when their clinicopathological profile is predictive of a good treatment response.^4-6^ This is critical in deciding approvals for unique subgroups, or conversely for discouraging use in the opposing subgroup – where negative biomarkers have demonstrated the lack of benefit. In addition, cost-effectiveness analysis (CEA), a major consideration in deciding against therapeutic strategies at a public health level, requires IPD data for the appropriate analysis. While new biomarkers for therapeutic regimens have been sought after and studied in trials, they remain exploratory and are often underpowered for interpretation. To overcome this, meta-analytic techniques may be exploited to pool data together to increase statistical power.

In an effort to enable secondary data analyses in such circumstances, we propose a workflow to retrieve unreported subgroup survival data from published KM curves.

## METHODS

*KMSubtraction* is an R-package designed to retrieve survival data of patients from unreported subgroups. An overview of the *KMSubtraction* workflow is illustrated in **Figure 1**. The workflow of *KMSubtraction* is detailed as such:

**Figure 1.**
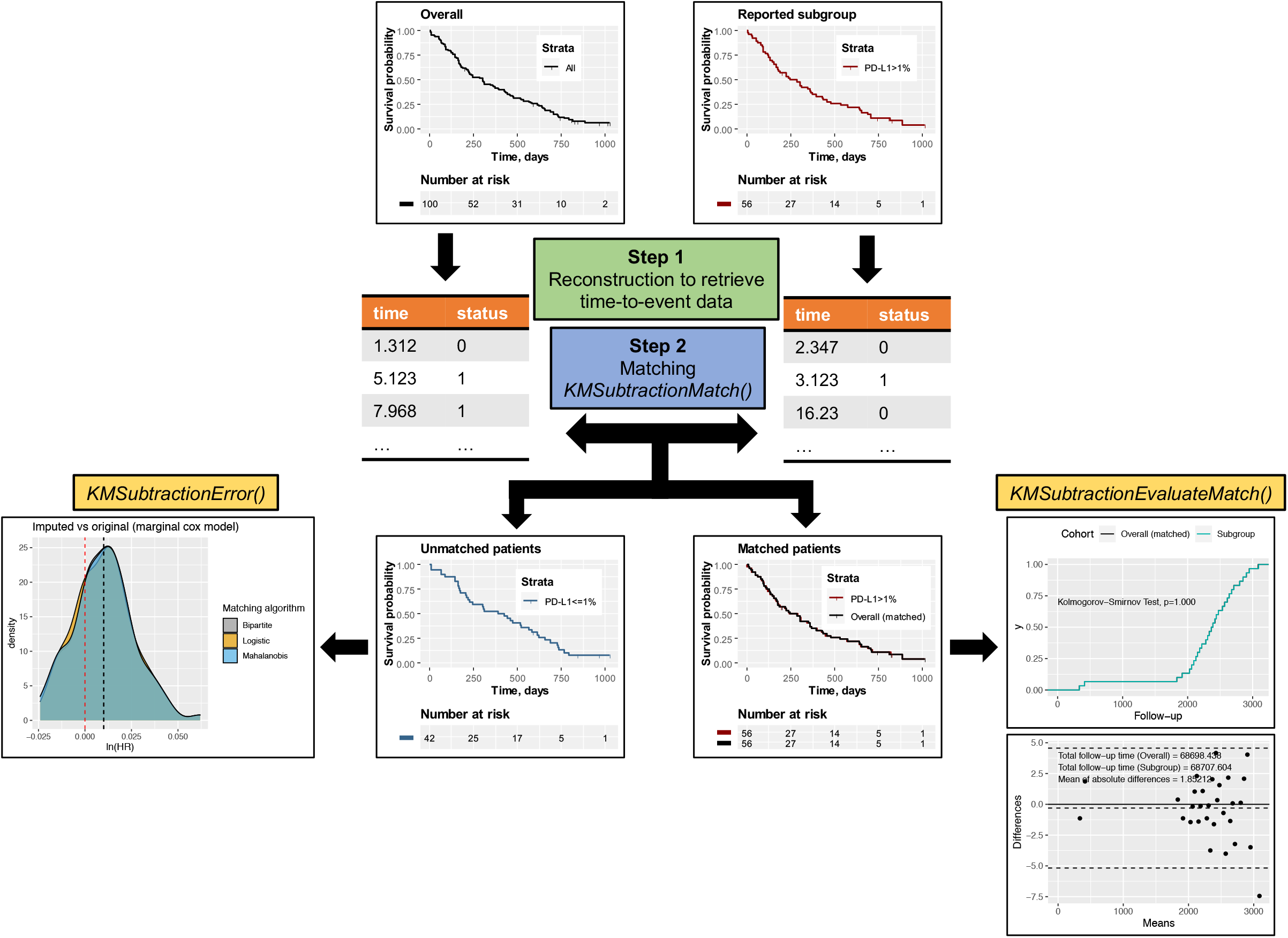
Illustration of *KMSubtraction*.

### Step 1 Reconstruction of time-to-event outcomes

A graphical reconstructive algorithm is exploited to estimate time-to-event outcomes from KM curves by an iterative algorithm based on KM estimation described by Guyot et al^1,2,7^. The implementation of this step is described by Liu et al in the package *IPDfromKM*^7^. KM curves describing the overall cohort and a known subgroup is required.

### Step 2 Matching of patients – *KMSubtractionMatch()*

*KMSubtractionMatch()* is a wrapper function that utilizing raw reconstructed time-to-event outcomes to match patients from subgroups among the overall cohort. Minimal cost bipartite matching with the Hungarian algorithm^8^ was adopted as the primary matching algorithm. The minimal cost bipartite matching algorithm aims to match patients from the overall and subgroup cohort by minimizing follow-up time differences between matched patients.^8^ The Hungarian alogrithm (or the Huhn-Munkres algorithm) is a combinatorial optimization algorithm that solves the problem of assigning optimal pairs in polynomial time, and is implemented in the *RcppHungarian*^9^ package in R. Matching through Mahalanobis distance matching (Mahalanobis) and nearest neighbor matching by logistic regression (Logisitic) with the *MatchIt*^10^ package are provided as well. Patients with events and censorships were matched separately. By excluding matched patients, the opposing unreported subgroup may be retrieved.

### Evaluation of matching – *KMSubtractionEvaluateMatch()*

The quality of matching was evaluated by comparing matched patients. This was conducted by inspecting the following parameters and quality of match test statistics (1) Bland-Altman^11^ plots to explore discrepancies between matched pairs with the *blandr*^12^ package in R (2) Empirical cumulative distribution functions and the Kolmogorov−Smirnov Test of follow-up times between matched pairs (3) KM curves of matched patients from the overall cohorts versus directly reconstructed subgroup cohorts, along with hazard ratios and log-rank tests from a marginal Cox proportional hazard model.

### Potential limits of error of each task – *KMSubtractionError()*

In light of the variation in each implementation, the limits of error surrounding each task would be different. It would therefore be of paramount importance to ascertain whether the implementation of *KMSubtraction* would be appropriate in each context. This may be especially relevant in the interpretation of derived data in situations where there is a sizable proportion of missing data in the opposing subgroup. *KMSubtractionError()* conducts Monte Carlo simulations to evaluate the limits of error of *KMSubtractionMatch()* given parameters surrounding the reconstruction task required. Follow-up time was modelled by a random weibull distribution of common shape parameter of 1.000 and scale parameter of 5.000. Reconstructed and original survival data were compared by means of marginal Cox-proportional hazard models and restricted mean survival time difference (RMST-D).^13^ The similarity may be summarized by the natural logarithmic of the hazard ratio, ln(HR) and RMST-D. Deviations from the true value is reflected by the absolute of ln(HR) and RMST-D, where |ln(HR)| and |RMST-D|=0 is ideal.

### Simulation exercise

Next, we conducted Monte Carlo simulations to evaluate the effect of the sample size, reported subgroup proportion, proportion of missing data, censorship proportion in the overall and subgroup cohort, and number of number-at-risk table intervals on the quality of reconstruction. 5,000 iterations were conducted for each scenario. Follow-up time was modelled by a random Weibull distribution of common shape parameter of 1.000 and scale parameter of 5.000. To facilitate the high magnitude of simulations performed, curve coordinates retrieval from KM curve images were automated by identifying RGB coordinates of each rasterized curve with a prespecified color and rescaling them with prespecified maximum and minimum xy coordinates. This was conducted using the *magick*^14^ package in R. The extracted xy coordinates of curves were thereafter incorporated into *IPDfromKM*^7^ for reconstruction. Utilizing *KMSubtraction*, patients from the opposing subgroup are identified.

Thereafter, reconstructed curves were compared against original curves using a marginal Cox proportional hazards model. The absolute value of natural logarithmic transformed hazard ratios (|ln(HR)|) was inspected as the main summary statistic. Running |ln(HR)| mean plots were inspected to evaluate for simulation convergence. For each scenario, means and Wald’s 95% confidence intervals were derived per outcome. The Pearson’s product-moment correlation coefficient r was inspected to evaluate the proportion of the variation in |ln(HR)| that is predictable from the upstream varying independent variables. Cutoffs were determined by the intersection between |ln(HR)|=0.03, 0.04 & 0.05 and smoothing splines of 15 degrees of freedom generated with the primary matching algorithm. Comparisons between the different matching algorithms were undertaken using two-way analysis of variance (ANOVA) with Tukey multiple pairwise-comparisons between each algorithm. Besides minimal cost bipartite matching, Mahalanobis distance matching, and nearest neighbor matching by logistic regression were explored as well. Finally, we sought to evaluate associations between the quality of match and the limits of error. Correlograms reflecting associations between |ln(HR)| and quality of match test statistics of *KMSubtractionEvaluateMatch()* were inspected to help interpret and identify tests that may assist in prognosticating reconstruction accuracy.

The R code for the simulations is provided in **Supplementary Code**, and the parameters utilized for each simulation scenario are provided in **Supplementary Table 1**. All analyses were conducted in R-4.1.0^15^.

## IMPLEMENTATION

A step-by-step user guide is provided in the **Supplementary Guide**.

### Scenario 1

The first scenario utilizes time-to-death data of patients with stage III colon carcinoma treated with fluorouracil (5FU) plus levamisole (Lev) versus levamisole only after resection in a randomized controlled trial by Moertel *et al*.^16^ The dataset was retrieved from the *survival* package in R.^17^ The hypothetical scenario is designed as such: KM curves describing survival outcomes for patients treated with Lev vs Lev+5FU is presented for the overall cohort (n=614) [**Figure 2A**] and for patients with more than 4 positive lymph nodes (n=168) [**Figure 2B**]. Survival outcomes for patients with less than or equal to 4 positive lymph nodes is however unreported (n=446) [**Figure 2C, solid line**].

**Figure 2.**
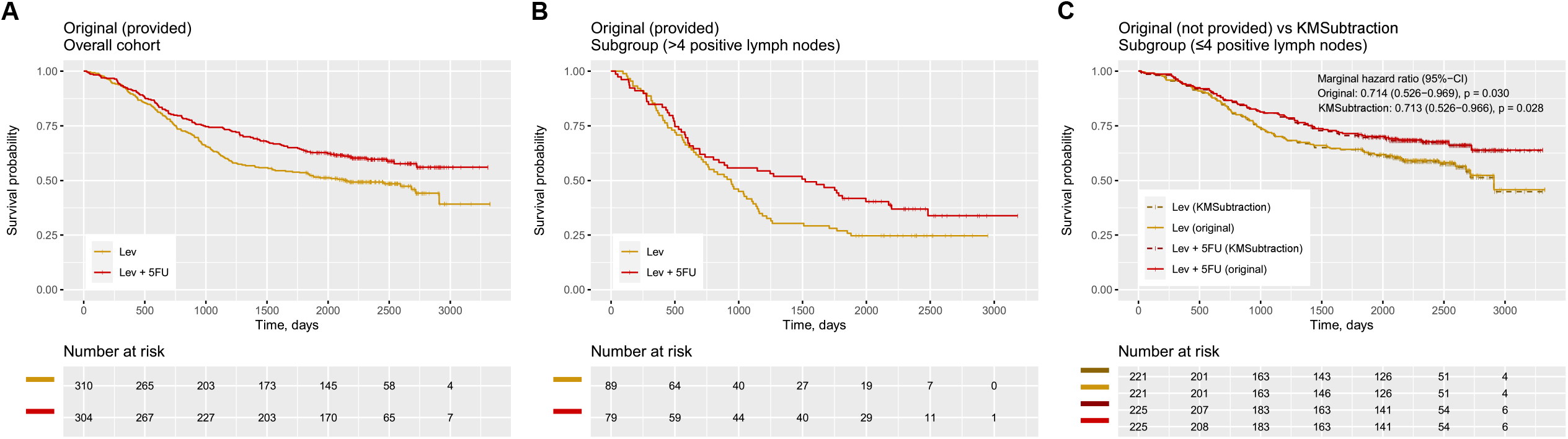
Example scenario 1 - patients with colon cancer treated with Levamisole (Lev) + 5-Fluorouracil (FU) vs Lev only. Kaplan Meier curves reporting (A) Overall cohort from ***original*** data (B) Patients with more than 4 positive lymph nodes (the presented subgroup) from ***original*** data (C) Patients with less than or equal to 4 positive lymph nodes comparing ***original and KMSubtraction derived*** data (opposing subgroup)

Upon reconstruction of **Figure 2A** and **Figure 2B**, and processing with *KMSubtractionMatch()* with minimal cost bipartite matching, the retrieved subgroup yielded curves [**Figure 2 C, dashed line**] similar to the original data. Hazard ratios on the marginal Cox model were similar as well (original-HR=0.714, 95%-CI: 0.526-0.969, p=0.030 vs *KMSubtraction*-HR=0.713, 95%-CI: 0.526-0.966, p=0.028) [**Figure 2C**].

Following, 1,000 Monte Carlo iterations with *KMSubtractionError()* were conducted per arm, yielding mean (standard deviation) |ln(HR)| of 0.00960 (0.00780) and 0.01120 (0.00866) for Lev and Lev+5FU respectively. This suggests that the limits of error of KMSubtraction in the above scenarios are likely small and negligible.

### Scenario 2

The second scenario utilizes time-to-death data of patients with primary breast cancer from the Rotterdam tumor bank by Royston and Altman *et al*.^18^ The dataset was retrieved from the *survival* package in R.^17^ The hypothetical scenario is designed as such: KM curves describing survival outcomes for patients treated with chemotherapy-only (Chemo) vs hormonal therapy-only (Hormon) is presented for the overall cohort (n=863) [**Figure 3A**] and for patients with estrogen receptors (ER) or progesterone receptors (PGR) more than 100 fmol/l (n=448) [**Figure 3B**]. Survival outcomes for patients with ER and PGR less than or equal to 100 fmol/l is however unreported (n=415) [**Figure 3C, solid line**].

**Figure 3.**
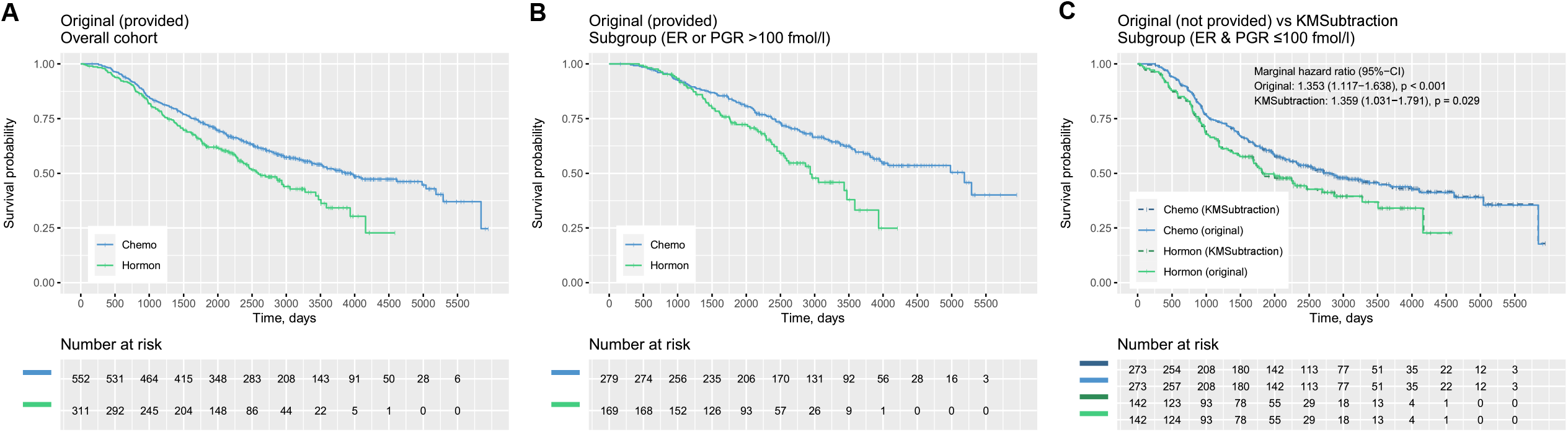
Example scenario 2 - patients with breast cancer treated with chemotherapy vs hormonal therapy. Kaplan Meier curves reporting (A) Overall cohort from ***original*** data (B) Patients with estrogen receptors (ER) or progesterone receptors (PGR) more than 100 fmol/l (the presented subgroup) from ***original*** data (C) Patients with less than or equal to 4 positive lymph nodes between ***original and KMSubtraction derived*** data (opposing subgroup)

Upon reconstruction of **Figure 3A** and **Figure 3B**, and processing with *KMSubtraction* with minimal cost bipartite matching, the retrieved subgroup yielded curves [**Figure 3C, dashed line**] similar to the original data. Hazard ratios on the marginal Cox model comparing Chemo vs Hormon among patients with ER and PGR less than or equal to 100 fmol/l were similar as well (original-HR=1.353, 95%-CI: 1.117-1.638, p<0.001 vs *KMSubtraction*-HR=1.359, 95%-CI: 1.031-1.791, p=0.029) [**Figure 3C**].

Following, 1,000 Monte Carlo iterations with *KMSubtractionError()* were conducted per arm, yielding mean (standard deviation) |ln(HR)| of 0.01211 (0.00818) and 0.01271 (0.01125) for Chemo and Hormon respectively. This suggests that the limits of error of KMSubtraction in the above scenarios are likely small and negligible.

## SIMULATION RESULTS

### Size of dataset

20 scenarios of intervals n=50 were created from sample sizes ranging 50 to 1000. |ln(HR)| was negatively associated with sample size (Bipartite: r=-0.374, p<0.001; Mahalanobis: r=-0.380, p<0.001; Logistic: r=-0.373, p<0.001), suggesting that small data sets are likely to yield high limits of error [**Figure 4A**]. No significant differences between the matching algorithms were demonstrated on the Tukey pairwise comparisons (all p>0.05). Datasets smaller than n=112, 89, 71 are likely to yield a mean |ln(HR)| of 0.03, 0.04 and 0.05 respectively with minimum cost bipartite matching. The mean of absolute differences in follow-up time among patients with events appears to be weakly associated with the limits of error [**Figure 5A**].

**Figure 4.**
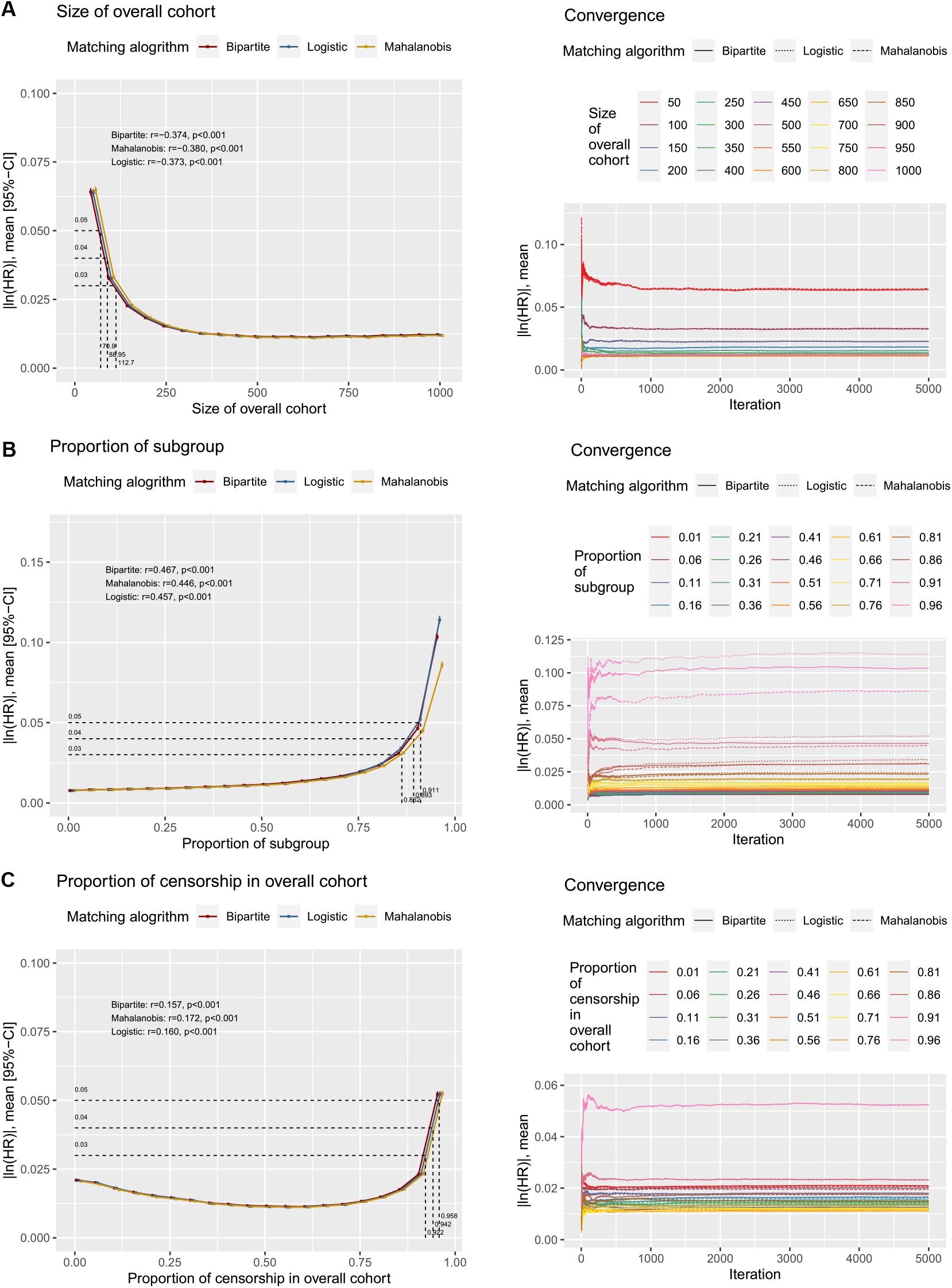
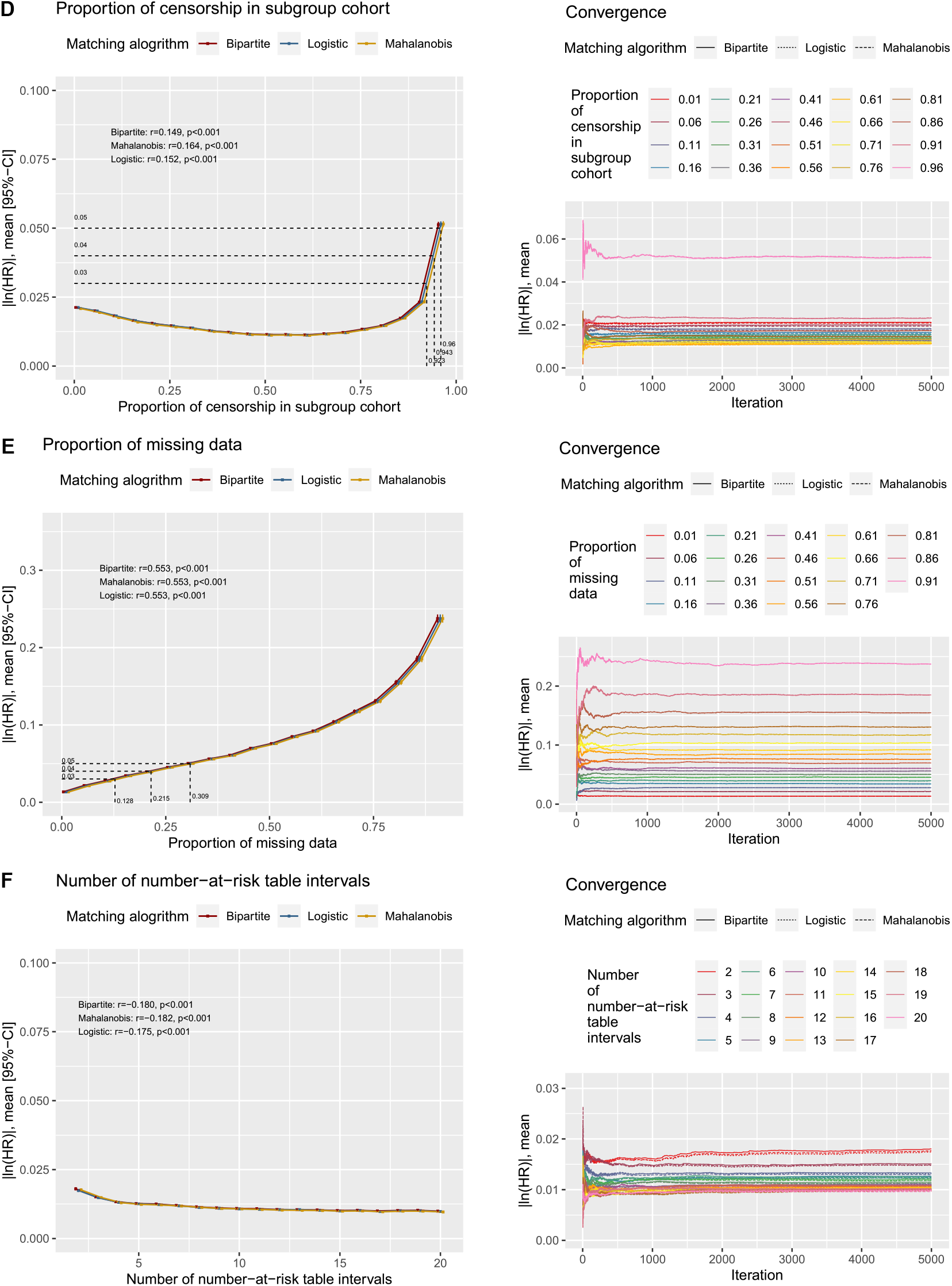
Outcomes of the simulation exercise and running mean plots demonstrating simulation convergence for (A) Size of dataset (B) Proportion of reported subgroup (C) Proportion of patients with censorship in the overall cohort (D) Proportion of patients with censorship in the subgroup cohort (E) Proportion of missing data (F) Number of number-at-risk table intervals

**Figure 5.**
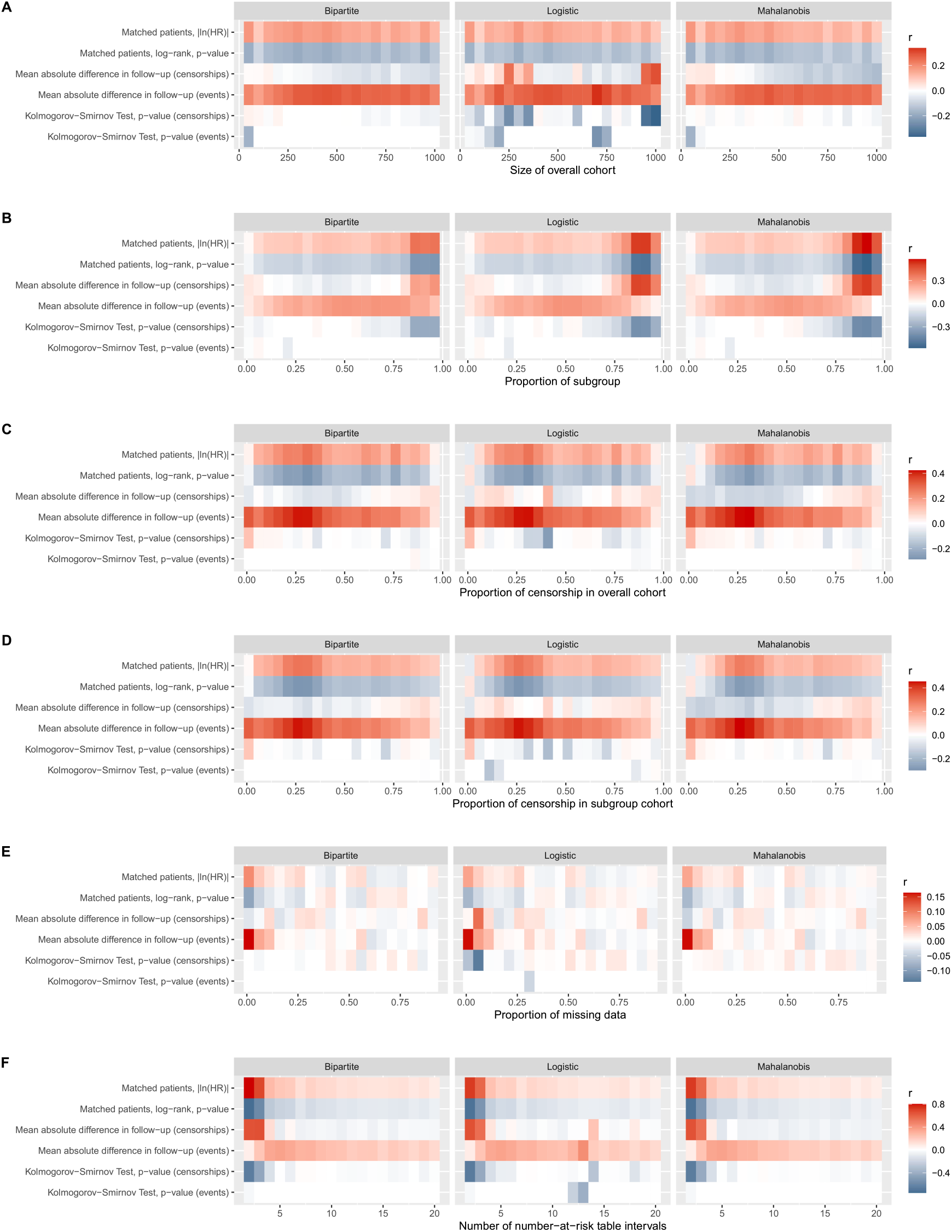
Correlograms of quality of match test statistics against the limits of error for simulations conducted for (A) Size of dataset (B) Proportion of reported subgroup (C) Proportion of patients with censorship in the overall cohort (D) Proportion of patients with censorship in the subgroup cohort (E) Proportion of missing data (F) Number of number-at-risk table intervals

### Proportion of reported subgroup

20 scenarios of intervals 5% were created from proportions ranging 1% to 96%. |ln(HR)| was positively associated with the proportion of reported subgroup (Bipartite: r=0.467, p<0.001; Mahalanobis: r=0.446, p<0.001; Logistic: r=0.457, p<0.01), suggesting that large proportion of reported subgroups are likely to yield high limits of error [**Figure 4B**]. The Mahalanobis matching algorithm yielded significantly lower limits of error when the proportion of reported subgroup is beyond 86% [**Supplementary Table 2**]. Proportions of reported subgroups larger than 86.2%, 89.3% and 91.1% are likely to yield a mean |ln(HR)| of 0.03, 0.04 and 0.05 respectively with minimum cost bipartite matching. In scenarios with high proportions of subgroups, the |ln(HR)| between matched patients of the overall and subgroup cohort, log-rank test and mean of absolute differences in follow-up time among patients with censorships appears to be associated with the limits of error [**Figure 5B**].

### Proportion of censorship in overall cohort

20 scenarios of intervals 5% were created from proportions ranging 1% to 96%. |ln(HR)| was weakly associated with the proportion of censorship (Bipartite: r=0.157, p<0.001; Mahalanobis: r=0.172, p<0.001; Logistic: r=0.160, p<0.001) [**Figure 4C**]. No significant differences between the matching algorithms were demonstrated on the Tukey pairwise comparisons (all p>0.05). Proportions of censorships in the overall cohort larger than 92.2%, 94.2% and 95.8% are likely to yield a mean |ln(HR)| of 0.03, 0.04 and 0.05 respectively with minimum cost bipartite matching. In scenarios with smaller proportions of censorship in the overall cohort, the mean of absolute differences in follow-up time among patients with events appears to be weakly associated with the limits of error [**Figure 5C**].

### Proportion of censorship in subgroup cohort

20 scenarios of intervals 5% were created from proportions ranging 1% to 96%. |ln(HR)| was weakly associated with the proportion of censorship (Bipartite: r=0.149, p<0.001; Mahalanobis: r=0.164, p<0.001; Logistic: r=0.152, p<0.001) [**Figure 4D**]. No significant differences between the matching algorithms were demonstrated on the Tukey pairwise comparisons (all p>0.05). Proportions of censorships in the overall cohort larger than 92.3%, 94.3% and 96.0% are likely to yield a mean |ln(HR)| of 0.03, 0.04 and 0.05 respectively with minimum cost bipartite matching. In scenarios with smaller proportions of censorship in the subgroup cohort, the mean of absolute differences in follow-up time among patients with events appears to be weakly associated with the limits of error [**Figure 5D**].

### Proportion of missing data

19 scenarios of intervals 5% were created from proportions ranging 1% to 91%. |ln(HR)| was positively associated with the proportion of reported subgroup (Bipartite: r=0.553, p<0.001; Mahalanobis: r=0.553, p<0.001; Logistic: r=0.553, p<0.001), suggesting that large proportion of missing data in unreported subgroups are likely to yield high limits of error [**Figure 4E**]. No significant differences between the matching algorithms were demonstrated on the Tukey pairwise comparisons (all p>0.05). Proportions of reported missing data larger than 12.8%, 21.5% and 30.9% are likely to yield a mean |ln(HR)| of 0.03, 0.04 and 0.05 respectively with minimum cost bipartite matching. Across the range of simulations, all quality of match test statistics were poorly associated the limits of error [**Figure 5E**].

### Number of number-at-risk table intervals

19 scenarios were created from intervals ranging from 1 to 20. |ln(HR)| was weakly associated with the number of number-at-risk table intervals (Bipartite: r=-0.180, p<0.001; Mahalanobis: r=-0.182, p<0.001; Logistic: r=-0.175, p<0.001) [**Figure 4F**].

No significant differences between the matching algorithms were demonstrated on the Tukey pairwise comparisons (all p>0.05). Across all scenarios, mean |ln(HR)| was under 0.03 with all matching algorithms. Where the number of number-at-risk table intervals were small, the |ln(HR)| between matched patients of the overall and subgroup cohort, log-rank test, mean of absolute differences in follow-up time among patients with censorships and the Kolmogorov-Smirnov Test of follow-up time between patients with censorships appears to be strongly associated with the limits of error [**Figure 5F**].

## DISCUSSION

Secondary analysis through meta-analysis has become increasingly relevant in this era of precision medicine, where definitive conclusions on biomarkers of disease or treatment regimens are required to facilitate clinical decision making. The upstream analyses demonstrates that *KMSubtraction* is robust and reliable.

The simulation exercise established that the limits of error of *KMSubtraction* were small and likely negligible in most circumstances. Apart from the proportion of missing data from the unreported subgroup, the reproducibility of reconstructed data was largely not affected by the other parameters studied. Interestingly, there was some evidence that Mahalanobis distance matching proffered a significantly smaller error when the proportion of the reported subgroup is beyond 86%. Nonetheless, given that *KMSubtraction* would be discouraged in scenarios where the proportion of reported subgroup is high, there is unlikely a scenario for this advantage to be exploited. There were otherwise no other appreciable differences on the Tukey HSD pairwise comparisons between the matching strategies investigated.

Extensive Monte Carlo simulations demonstrate that datasets with high reported subgroup proportion (r=0.467, p<0.001), small dataset size (r=-0.374, p<0.001) and high proportion of missing data in the unreported subgroup (r=0.553, p<0.001) were associated with uncertainty are likely to yield high limits of error with *KMSubtraction*. Users are advised against implementation of *KMSubtraction* in such situations, or if need be, transparently acknowledge the ensuing limitations and likely compromise in quality of reconstructed data. The limits of error of *KMSubtraction*, as reflected by the mean |ln(HR)| from converged Monte Carlo simulations may be arbitrarily interpreted as small (<0.03), moderate (0.03-0.05) and large (>0.05), and may guide the appropriateness of implementing *KMSubtraction* in each context.

In implementing *KMSubtraction*, the quality of match may be the only raw parameter derived from the manually retrieved survival information. From our simulations, we found a wide variation of how the quality of match influences the limits of error of *KMSubtraction*. As such, in circumstances where the simulated limit of error is shown to be high, users are advised to inspect the association of each quality of match test statistic against the simulated limits of error.

This approach is not without its limitations. The described method is also unable to retrieve patient-level covariates for adjustment. Hence, biomarker investigation through this analysis should be interpreted with caution as no further analysis of causal inference may be conducted. Further, the described method is only able to handle dichotomous variables, rather than continuous variables. This would unfortunately limit meta-analytic pooling of unreported subgroup data after *KMSubtraction* when different cutoffs are used instead.

Given the demonstrated effect of the above parameters on the quality of *KMSubtraction* derived data, it would be instructive for those utilizing *KMSubtraction* to report (1) Proportion of reported subgroup among the overall cohort (2) Size of dataset (3) Proportion of censorship in overall and subgroup cohorts (4) Proportion of missing data (5) Number of number-at-risk table intervals (6) Mean and standard deviation of |ln(HR)| from *KMSubtractionError()* simulations.

### Conclusion

While *KMSubtraction* demonstrates robustness in deriving survival data from unreported subgroups, the implementation of *KMSubtraction* should take into consideration the aforementioned limitations. The limits of error of *KMSubtraction*, as reflected by the mean |ln(HR)| from converged Monte Carlo simulations may guide the interpretation of reconstructed survival data of unreported subgroups. This approach hopes to enhance the quality of secondary analysis of subgroups and biomarkers in the era of precision medicine.

## Data Availability

Not applicable, the data was derived from simulations.

## Acknowledgements

The authors would like to thank Assistant Professor Justin Silverman, College of Information Science and Technology, Penn State University, USA, for clarifying enquiries regarding the use of the Hungarian algorithm for bipartite matching; and Associate Professor Hyungwon Choi, Department of Medicine, Yong Loo Lin School of Medicine, National University of Singapore, Singapore, for guiding us through the simulations and development of the package.

