## Supplementary Materials for "KMSubtraction: Reconstruction of unreported subgroup survival data utilizing published Kaplan-Meier survival curves"

**Supplementary Guide**

The vignette is made available on github: [https://github.com/josephjzhao/KMSubtraction](https://github.com/josephjzhao/KMSubtraction#readme)

**Supplementary Table 1** Simulation parameters

The independent variable (denoted by red boxes) follows according to the following arithmetic sequence: $\lim_{n\to m} (a+(1-n)*b)$

| **Variable being investigated** | **Size of overall cohort (*N*_overall_)** | **Proportion of reported subgroup (*P*_subgroup_)** | **Proportion of patients with censorship in overall cohort**  **(*P*_overall, censored_)** | **Proportion of patients with censorship in subgroup cohort**  **(*P*_subgroup, censored_)** | **Proportion of missing data (*P*_missing_)** | **Number of number-at-risk table intervals**  **(*N*_interval_)** | **Number of Monte Carlo iterations**  **(i)** |
| --- | --- | --- | --- | --- | --- | --- | --- |
| ***N*_overall_** | $a=50,$  $b=1000,$  $m=20$ | 0.50 | 0.50 | 0.50 | 0.00 | 8 | 5000 |
| ***P*_subgroup_** | 500 | $a=0.01,$  $b=0.05,$  $m=20$ | 0.50 | 0.50 | 0.00 | 8 | 5000 |
| ***P*_overall, censored_** | 500 | 0.50 | $a=0.01,$  $b=0.05,$  $m=20$ | * | 0.00 | 8 | 5000 |
| ***P*_subgroup, censored_** | 500 | 0.50 | * | $a=0.01,$  $b=0.05,$  $m=20$ | 0.00 | 8 | 5000 |
| ***P*_missing_** | 500 | 0.50 | 0.50 | 0.50 | $a=0.01,$  $b=0.05,$  $m=19$ | 8 | 5000 |
| ***N*_interval_** | 500 | 0.50 | 0.50 | 0.50 | 0.00 | $a=2,$  $b=1,$  $m=19$ | 5000 |

* Since subgroups are dichotomous, the relationship between *P*_overall, censored_ and *P*_subgroup, censored_ is bounded by

$\frac{Psubgroup*Poverall, censored}{Psubgroup, censored}=\frac{Nsubgroup, censored}{Noverall, censored}\leq1$

_Thus,_ when the proportion of censored patients from the overall and subgroup cohorts were investigated as the independent variable, we arbitrarily defined $\frac{Nsubgroup, censored}{Noverall, censored}=0.5$_._

This gives $Psubgroup, censored=\frac{P\mathrm{subgroup}*Poverall, censored}{0.5}$ _and_ $Poverall, censored=\frac{0.5*Psubgroup, censored}{P\mathrm{subgroup}}$

when the proportion of censored patients from the overall and subgroup cohorts were investigated respectively.

**Supplementary Table 2** Significant outcomes of Tukey multiple pairwise-comparisons between each matching algorithm

| **Comparison of matching algorithms** | **Parameters** | | | | | | **Difference (95% CI)** | **p** |
| --- | --- | --- | --- | --- | --- | --- | --- | --- |
|  | **Size of overall cohort (*N*_overall_)** | **Proportion of reported subgroup (*P*_subgroup_)** | **Proportion of patients with censorship in overall cohort**  **(*P*_overall, censored_)** | **Proportion of patients with censorship in subgroup cohort**  **(*P*_subgroup, censored_)** | **Proportion of missing data (*P*_missing_)** | **Number of number-at-risk table intervals**  **(*N*_interval_)** |  |  |
| **Logistic/Bipartite** | 500 | 0.86 | 0.50 | 0.50 | 0.00 | 8 | 0.003166297 (0.001300484 to 0.005032110) | **<0.001** |
| **Mahalanobis/Logistic** | 500 | 0.86 | 0.50 | 0.50 | 0.00 | 8 | -0.002886512 (-0.004752325 to -0.001020700) | **<0.001** |
| **Logistic/Bipartite** | 500 | 0.91 | 0.50 | 0.50 | 0.00 | 8 | 0.005700769 ( 0.003834957 to 0.007566582) | **<0.001** |
| **Mahalanobis/Logistic** | 500 | 0.91 | 0.50 | 0.50 | 0.00 | 8 | -0.007242428 (-0.009108240 to -0.005376615) | **<0.001** |
| **Logistic/Bipartite** | 500 | 0.96 | 0.50 | 0.50 | 0.00 | 8 | 0.010563824 (0.008698011 to 0.012429636) | **<0.001** |
| **Mahalanobis/Bipartite** | 500 | 0.96 | 0.50 | 0.50 | 0.00 | 8 | -0.017536851 (-0.019402664 to -0.015671039) | **<0.001** |
| **Mahalanobis/Logistic** | 500 | 0.96 | 0.50 | 0.50 | 0.00 | 8 | -0.028100675 (-0.029966488 to -0.026234863) | **<0.001** |

Abbreviations: Logistic, nearest neighbor matching with distances defined by logistic regression; Bipartite, minimal cost bipartite matching; Mahalanobis, Mahalanobis distance matching; CI, confidence interval.

**Supplementary Code**

\# Implemented in R-4.1.0

main_wd="C:/Users/jzhao/OneDrive/Research_Cloud/NUH_NCIS/KMSubtraction/"

setwd(main_wd)

\# number of Monte Carlo iterations per scenario

n.mc=5000

\# set number of cores

detectCores()

n.cores=6

\#### DOWNLOAD libraries ####

package.name=c(# unique to KMSubtraction

"MatchIt", "wakefield", "magick", "RcppHungarian", "scales", "IPDfromKM", "svglite", "rsvg", "blandr", "KMSubtraction",

\# data manipulation

"tidyverse", "reshape", "readr", "readxl", "dplyr", "tidyr", "lubridate", "tibble", "plyr", "devtools", "stringr", "stringi",

\# parallel processing

"doParallel", "parallel", "foreach", "doSNOW",

\# survival analysis

"survminer", "survival",

\# misc

"cluster", "ResourceSelection", "progress"

)

for (package.name in package.name){

if (!require(package.name, character.only = TRUE)){

install.packages(package.name, character.only = TRUE)

library(package.name, character.only = TRUE)} else {library(package.name, character.only = TRUE)}}

\#### LOAD functions ####

seq_sim=function(x){seq(from=as.numeric(str_extract(x, "^(\\d|\\.)*(?=;)")),

to=as.numeric(str_extract(x, "(?<=;)(\\d|\\.)*(?=;)")),

as.numeric(str_extract(x, "(?<=;)(\\d|\\.)*$")))}

\#### LOAD Simulation parameters ####

simulation_parameters=data.frame(outcome = c("outcome_size", "outcome_subgroup", "outcome_censorship_overall", "outcome_censorship_subgroup", "outcome_missing", "outcome_interval"),

n = c("50;1000;50", "500;500;0", "500;500;0", "500;500;0", "500;500;0", "500;500;0"),

subgroup.p = c("0.5;0.5;0", "0.01;0.99;0.05", "0.5;0.5;0", "0.5;0.5;0", "0.5;0.5;0", "0.5;0.5;0"),

censor_overall.p = c("0.5;0.5;0", "0.5;0.5;0", "0.01;0.99;0.05","0.5;0.5;0", "0.5;0.5;0", "0.5;0.5;0"),

censor_subgroup.p = c("0.5;0.5;0", "0.5;0.5;0", "0.5;0.5;0", "0.01;0.99;0.05", "0.5;0.5;0", "0.5;0.5;0"),

missing.p = c("0;0;0", "0;0;0", "0;0;0", "0;0;0", "0.01;0.91;0.05", "0;0;0"),

interval = c("8;8;0", "8;8;0", "8;8;0", "8;8;0", "8;8;0", "2;20;1"),

mc = rep(paste0("1;", n.mc,";1" ), 6))

\# count the total number of iterations

iterations=0

for (r in 1:nrow(simulation_parameters)){

row.iterations=1

for (c in 2:ncol(simulation_parameters)){

row.iterations=row.iterations*length(seq_sim(simulation_parameters[r,c]))

}

iterations=row.iterations+iterations

}

print(paste0("Total number of iterations: ", iterations))

set.seed(NULL)

\#### Prepare parallel processing settings ####

registerDoParallel(n.cores)

registerDoSNOW(makeSOCKcluster(n.cores))

\# progress bar

pb <- txtProgressBar(min=1, max=iterations, style=3)

progress <- function(n) setTxtProgressBar(pb, n)

opts <- list(progress=progress)

\#### Simulation ####

df_out=NULL

i.outcome=1

system.time({

df_out=

foreach (i.outcome = 1:length(simulation_parameters$outcome), .options.snow=opts, .combine=rbind) %:%

foreach (i.n = 1:length(seq_sim(simulation_parameters$n[i.outcome])), .options.snow=opts, .combine=rbind) %:%

foreach (i.censor_overall = 1:length(seq_sim(simulation_parameters$censor_overall.p[i.outcome])), .options.snow=opts, .combine=rbind) %:%

foreach (i.censor_subgroup = 1:length(seq_sim(simulation_parameters$censor_subgroup.p[i.outcome])), .options.snow=opts, .combine=rbind) %:%

foreach (i.subgroup = 1:length(seq_sim(simulation_parameters$subgroup.p[i.outcome])), .options.snow=opts, .combine=rbind) %:%

foreach (i.missing = 1:length(seq_sim(simulation_parameters$missing.p[i.outcome])), .options.snow=opts, .combine=rbind) %:%

foreach (i.interval = 1:length(seq_sim(simulation_parameters$interval[i.outcome])), .options.snow=opts, .combine=rbind) %:%

foreach (i.mc = 1:length(seq_sim(simulation_parameters$mc[i.outcome])), .options.snow=opts, .combine=rbind) %dopar% {

library(survival)

library(survminer)

library(wakefield)

library(svglite)

library(magick)

library(stringi)

library(stringr)

library(dplyr)

library(tidyr)

library(tidyverse)

library(scales)

library(svglite)

library(rsvg)

library(IPDfromKM)

library(readr)

library(RcppHungarian)

library(dplyr)

library(MatchIt)

library(survRM2)

library(blandr)

library(KMSubtraction)

\#### 0_Define parameters ####

outcome=simulation_parameters$outcome[i.outcome]

\# assumptions and settings

mc=seq_sim(simulation_parameters$mc[i.outcome])

n=seq_sim(simulation_parameters$n[i.outcome])

censor_overall.p=seq_sim(simulation_parameters$censor_overall.p[i.outcome])

censor_subgroup.p=seq_sim(simulation_parameters$censor_subgroup.p[i.outcome])

missing.p=seq_sim(simulation_parameters$missing.p[i.outcome])

subgroup.p=seq_sim(simulation_parameters$subgroup.p[i.outcome])

interval=seq_sim(simulation_parameters$interval[i.outcome])

\#### 1_Create simulation data ####

if (outcome=="outcome_censorship_overall"){

censor_subgroup.p[i.censor_subgroup]=(subgroup.p[i.subgroup]*censor_overall.p[i.censor_overall])/0.5

}

if (outcome=="outcome_censorship_subgroup"){

censor_overall.p[i.censor_overall]=(0.5*censor_subgroup.p[i.censor_subgroup])/(subgroup.p[i.subgroup])

}

\# number of patients per subgroup

n.subgroup=round(subgroup.p[i.subgroup]*n[i.n],0)

n.opposingsubgroup=round((n[i.n]-n.subgroup),0)

\# number of censored patients in the opposing subgroup

n.censoroverall=round(censor_overall.p[i.censor_overall]*n[i.n], 0)

n.censorsubgroup=round(censor_subgroup.p[i.censor_subgroup]*n.subgroup,0)

n.censoroppsingsubgroup=n.censoroverall-n.censorsubgroup

censor_opposingsubgroup.p=n.censoroppsingsubgroup/n.opposingsubgroup

\# generating simulated data

df_overall=NULL

df_overall$time=rweibull(n.subgroup+n.opposingsubgroup, shape=1,scale=5)

df_overall$status=c(rep(0, n.censorsubgroup),

rep(1, n.subgroup-n.censorsubgroup),

rep(0, n.censoroppsingsubgroup),

rep(1, n.opposingsubgroup-n.censoroppsingsubgroup))

df_overall$subgroup=c(rep(1, n.subgroup),

rep(0, n.opposingsubgroup))

df_overall$subgroup[df_overall$subgroup==0]=r_sample_binary(length(df_overall$subgroup[df_overall$subgroup==0]), x = c(NA,0), prob = c(missing.p[i.missing], 1-missing.p[i.missing]), name = "Binary")

df_overall=data.frame(df_overall)

df_subgroup=subset(df_overall, df_overall$subgroup==1)

group=paste0("_n",n[i.n],

"_subgroup", subgroup.p[i.subgroup],

"_censoroverall", censor_overall.p[i.censor_overall],

"_censorsubgroup", censor_subgroup.p[i.censor_subgroup],

"_missing", missing.p[i.missing],

"_interval", interval[i.interval],

"_mc", mc[i.mc])

if(nrow(df_subgroup)==0){

stop("Subgroup=0")

}

km_overall=survfit(Surv(time, status) ~ 1, data=df_overall)

km_subgroup=survfit(Surv(time, status) ~ 1, data=df_subgroup)

df_overall_clicks=getxycoordinates(df_overall, label=paste0("overall",group))

df_subgroup_clicks=getxycoordinates(df_subgroup, label=paste0("subgroup",group))

\#### 2_Reconstruction ####

\# at risk tables

df_overall_risktable=ggsurvtable(km_overall, data=df_overall, break.time.by = (max(df_overall$time)/interval[i.interval]))$risk.table$data[,2:3]

df_subgroup_risktable=ggsurvtable(km_subgroup, data=df_subgroup, break.time.by = (max(df_subgroup$time)/interval[i.interval]))$risk.table$data[,2:3]

\# overall

df_overall_recon=getIPD(prep=preprocess(dat=df_overall_clicks,

trisk=df_overall_risktable$time,

nrisk=df_overall_risktable$n.risk,

maxy=1),

armID=1, tot.events=NULL)$IPD

\# subgroup

df_subgroup_recon=getIPD(prep=preprocess(dat=df_subgroup_clicks,

trisk=df_subgroup_risktable$time,

nrisk=df_subgroup_risktable$n.risk,

maxy=1),

armID=1, tot.events=NULL)$IPD

\#### 3_Matching ####

match.algo=c("bipartite", "maha", "logit")

tbl=NULL

for (i.match in match.algo){

df_match=KMSubtractionMatch(df_overall_recon, df_subgroup_recon, matching=i.match)$data

df_match$strata="matched"

df_overall$strata="original"

df_combined=bind_rows(df_match, df_overall)

\# survival analysis

sum.cox=coxph(formula = Surv(time, status) ~ strata, data=subset(df_combined, df_combined$subgroup==0)) %>% summary

GT=coxph(formula = Surv(time, status) ~ strata, data=subset(df_combined, df_combined$subgroup==0)) %>% cox.zph

\# collate

tbl=rbind(tbl,c(outcome=outcome,

n=n[i.n],

subgroup.p=subgroup.p[i.subgroup],

censorship_overall.p=censor_overall.p[i.censor_overall],

censorship_subgroup.p=censor_subgroup.p[i.censor_subgroup],

missing.p=missing.p[i.missing],

interval=interval[i.interval],

mc=mc[i.mc],

logrank=sum.cox$sctest[3],

HR=sum.cox$coefficients[2],

TE=sum.cox$coefficients[1],

se=sum.cox$coefficients[3],

GT.p=GT$table[1,3],

matching=i.match))

}

\#### 4_Export ####

tbl

}

})

\# clear parallel processing

stopImplicitCluster()

close(pb)

stopCluster(makeSOCKcluster(n.cores))

\#### Export as csv ####

\# cleaning

df_out=data.frame(df_out)

df_out[,-grep("matching|outcome", colnames(df_out))]=apply(df_out[,-grep("matching|outcome", colnames(df_out))],2, as.numeric)

\# write

write.csv(df_out, paste0("df_run.csv"))
